# Acute COVID-19 gene-expression profiles show multiple etiologies of long-term sequelae

**DOI:** 10.1101/2021.10.04.21264434

**Authors:** Ryan C. Thompson, Nicole W. Simons, Lillian Wilkins, Esther Cheng, Diane Marie Del Valle, Gabriel E. Hoffman, Brian Fennessy, Konstantinos Mouskas, Nancy J. Francoeur, Jessica S. Johnson, Lauren Lepow, Jessica Le Berichel, Christie Chang, Aviva G. Beckmann, Ying-chih Wang, Kai Nie, Nicholas Zaki, Kevin Tuballes, Vanessa Barcessat, Mario A. Cedillo, Laura Huckins, Panagiotis Roussos, Thomas U. Marron, The Mount Sinai COVID-19 Biobank Team, Benjamin S. Glicksberg, Girish Nadkarni, Edgar Gonzalez-Kozlova, Seunghee Kim-Schulze, Robert Sebra, Miriam Merad, Sacha Gnjatic, Eric E. Schadt, Alexander W. Charney, Noam D. Beckmann

## Abstract

Two years into the SARS-CoV-2 pandemic, the post-acute sequelae of infection are compounding the global health crisis. Often debilitating, these sequelae are clinically heterogeneous and of unknown molecular etiology. Here, a transcriptome-wide investigation of this new condition was performed in a large cohort of acutely infected patients followed clinically into the post-acute period. Gene expression signatures of post-acute sequelae were already present in whole blood during the acute phase of infection, with both innate and adaptive immune cells involved. Plasma cells stood out as driving at least two distinct clusters of sequelae, one largely dependent on circulating antibodies against the SARS-CoV-2 spike protein and the other antibody-independent. Altogether, multiple etiologies of post-acute sequelae were found concomitant with SARS-CoV-2 infection, directly linking the emergence of these sequelae with the host response to the virus.

## Introduction

Since the outbreak of severe acute respiratory syndrome coronavirus 2 (SARS-CoV-2), over 232 million individuals have developed coronavirus disease 2019 (COVID-19) worldwide. The post-acute sequelae of SARS-CoV-2 infection (PASC) comprise a broad array of symptoms that emerge after recovery in up to 30% of individuals, and over 40% of those requiring hospitalization^1,2^. These symptoms include, among others, overall fatigue, dyspnea, smell and taste problems, and often last over long periods of time^3-6^. Elements of the immune response during the acute phase of COVID-19 have been shown to be associated with certain PASC outcomes^6-10^. However, sample sizes to date have been small and the scope of molecular profiling limited. There is therefore a need to comprehensively characterize the molecular processes occurring upon infection that associate with subsequent development of PASC.

Here, whole-blood gene expression and antibody titers were profiled in a large cohort of hospitalized COVID-19 patients followed clinically into the post-acute period^11^. Multiple distinct acute phase gene expression signatures were identified, linking several immune cell types to post-acute sequelae. At least two independent etiologies of PASC were found, distinguished by their dependence on the titers of antibodies against the SARS-CoV-2 spike surface protein. Together, our results show that the molecular processes leading to PASC are already detectable during the acute phase of COVID-19, establish multiple distinct etiologies leading to different long-term outcomes, and directly link the emergence of these symptoms to the host-response to SARS-CoV-2 infection.

## Results

### PASC characteristics in study cohort

The Mount Sinai COVID-19 Biobank is an ongoing study within the Mount Sinai Health System in New York City. As outlined in Fig. 1, the current report is limited to 567 subjects (495 hospitalized with COVID-19 and 72 healthy and hospitalized controls) enrolled between April and June of 2020 (Supplementary Table 1). Blood was collected from hospitalized subjects throughout their hospital stay and from healthy controls at a single time point in the outpatient setting (1392 total samples). Six months or more after discharge, 251 subjects completed a self-report checklist assessing for the emergence of PASC (Fig. 2a, b). No symptoms were significantly more frequent in recovered COVID-19 cases relative to formerly hospitalized controls, potentially due to the small number of controls that completed the PASC checklist (N = 15, Supplementary Table 1). Within cases, maximum COVID-19 severity and any intensive care unit (ICU) encounter were not significantly associated with symptoms (Extended data Fig. 1). Among other prior comorbidities, laboratory values and medications, there were 8 significant associations with PASC symptoms out of 2780 tests (Supplementary Table 1). These associations were not consistent across symptoms. Additionally, anti-spike antibody titers were only associated with sleep problems (Supplementary Table 1). Finally, significant co-occurrence between some pairs of symptoms, but not others, were observed, with the appearance of at least two distinct clusters, related respectively to lung and neuropsychiatric symptoms (Fig. 2c, Extended Data Fig. 1).

**Figure 1:**
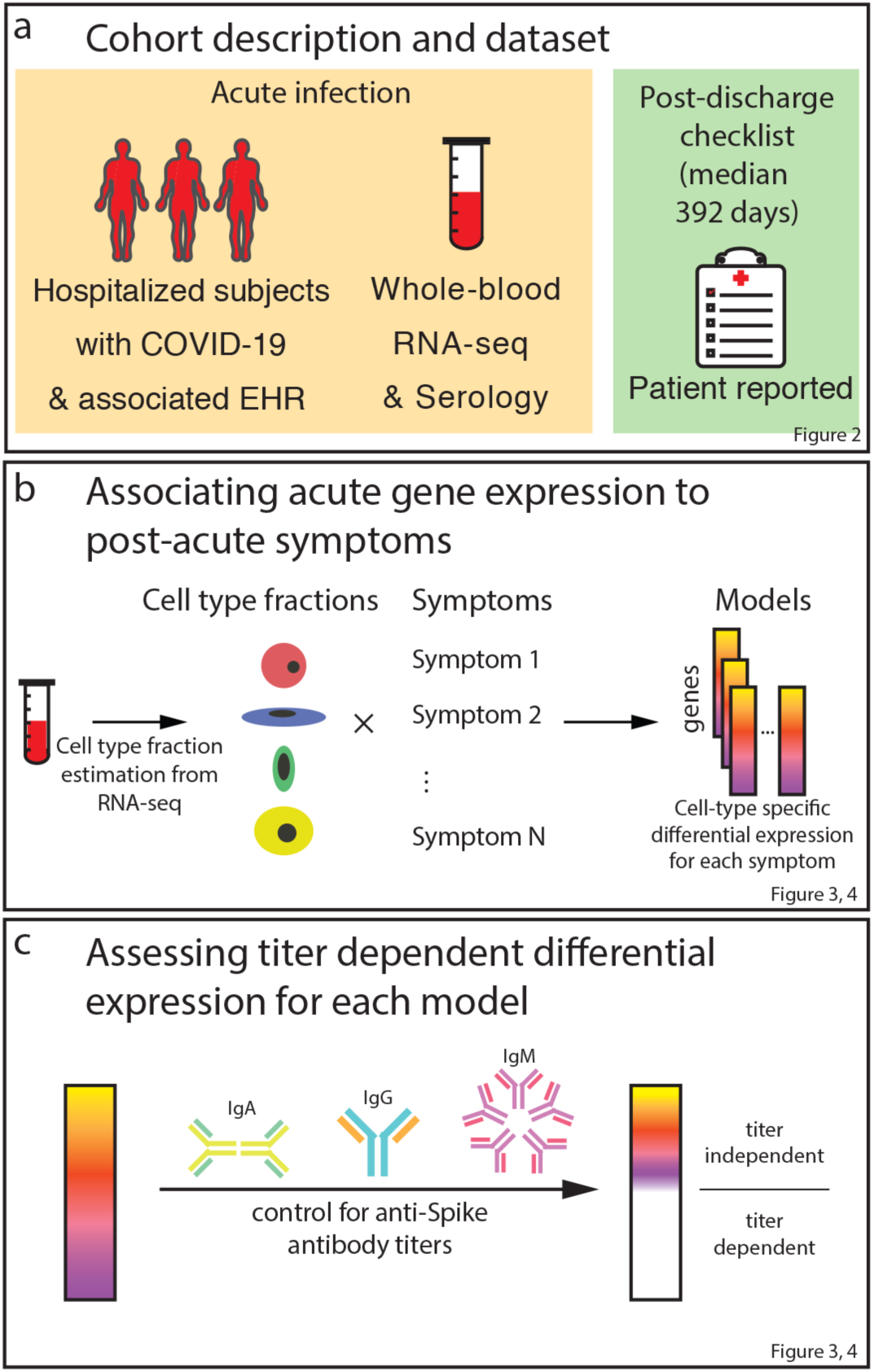
Study workflow.

**Figure 2:**
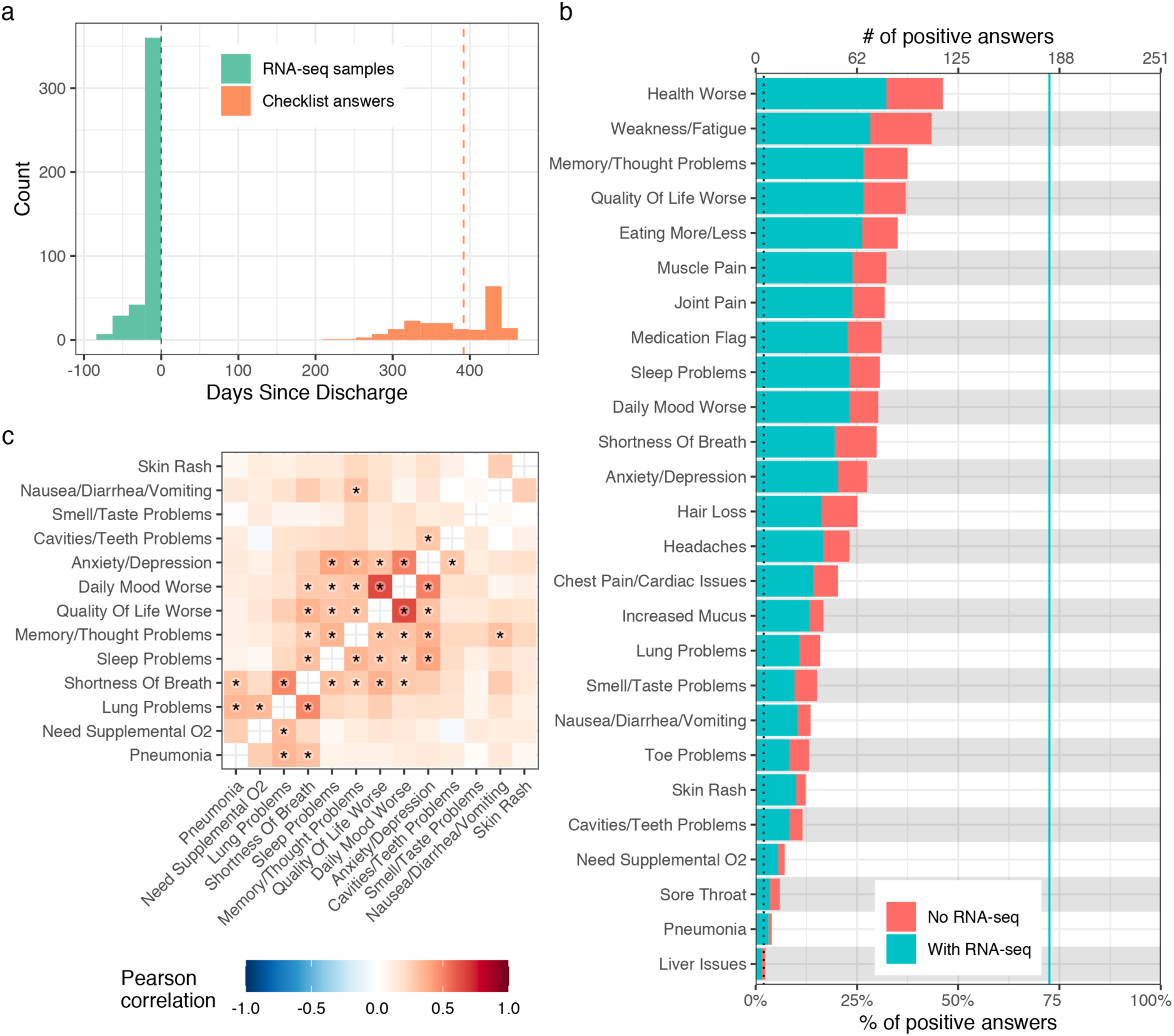
Description of PASC checklist items. a) Histogram of the timing of blood sampling and PASC checklist completion. The x and y axes are the number of days since discharge and a count of observations respectively. The green bars are counts of RNA-seq samples and orange bars the number of days between COVID-19 hospitalization discharge (black dashed line) and PASC checklist completion (dashed orange line is the median). b) Prevalence of PASC checklist items in our cohort. The y axis is symptoms, the upper and lower x axes are the number of positive answers and percentage of subjects from the entire cohort with a positive answer, respectively. The blue line represents the subset of subjects with RNA-seq who completed the checklist. The dashed black line is the cutoff used for inclusion in follow up analyses. c) PASC checklist item correlations. The axes are representative of the symptoms of interest (Methods) and the color the Pearson correlation of their coincidence. Correlations with family wise error rate (FWER, Holm 1979) adjusted p values < 0.05 are indicated with a star. Rows and columns are ordered to minimize distance between adjacent symptoms.

### Cell type specific differential expression

We hypothesized that the relationship between the acute phase of COVID-19 and the development of post-acute sequelae is reflected in blood gene expression. RNA sequencing (RNA-seq) was performed in 438 acute-phase samples from 182 of the 251 subjects that had completed the PASC checklist. All genes were tested for association with each symptom in the checklist, accounting for any ICU encounter and severity at the time of sampling. In addition, cell-type specific changes in gene expression were assessed for association with each symptom. To do this from whole-blood expression data, cell-type fractions were first estimated using standard bioinformatic approaches and validated using complete blood counts (Extended Data Fig. 2a). Higher plasma cell and lower follicular helper T cell fractions were associated with post-acute pneumonia and muscle pain respectively, but most PASC symptoms had no significant associations with any cell-type fractions (Supplementary Table 1). To test whether a gene’s cell-type-specific expression was different between subjects with and without a symptom, a differential expression model was fit with an interaction term. This model was fit for all cell-types where at least one percent of the variation in estimated fractions was explained by COVID-19 severity (Extended Data Fig. 2b). The cell-type-specific expression encompasses both the gene’s expression in that cell type and outside of that cell type but correlated to its relative fraction (Extended Data Fig. 2c). For example, a gene expressed in cell type A that differentially regulates proliferation of cell type B would be significant in the interaction model for cell type B even though it is not expressed in that cell type. To verify that our models defined cell-type-specific differential expression (DE) as expected, differentially expressed genes (DEGs) were compared to the corresponding cell-type specific markers from the literature and a significant overlap was found in the majority of instances (Supplementary Table 2). Our study primarily focused on cell-types whose DEGs were enriched for its own markers.

After accounting for blood composition, no DEGs were found in whole blood for any symptoms (Extended Data Fig. 3, Supplementary Table 3). In contrast, many symptoms showed significant DE in cell-type-specific tests (Fig. 3a, b, red bars) and their respective signatures were further annotated for known biology using gene ontology (GO) term enrichment analysis (Fig. 3c, Supplementary Table 4, Extended Data Fig. 4). Genes with significantly higher and lower expression in patients with a symptom are hereafter referred to as upregulated and downregulated, respectively. Plasma cells had the largest number of symptom-associated DE signatures with at least one hundred DEGs: sleep problems, lung problems, nausea/diarrhea/vomiting, skin rash, smell/taste problems, and pneumonia. Notably, the DEGs for pneumonia were almost entirely downregulated, not simply recapitulating the association with higher plasma cell fraction. CD8^+^ and γδ T cells were associated with a worse quality of life; memory resting CD4^+^ T cells and neutrophils were associated with cavities/teeth problems; and memory activated CD4^+^ T cells were associated with memory/thought problems.

**Figure 3:**
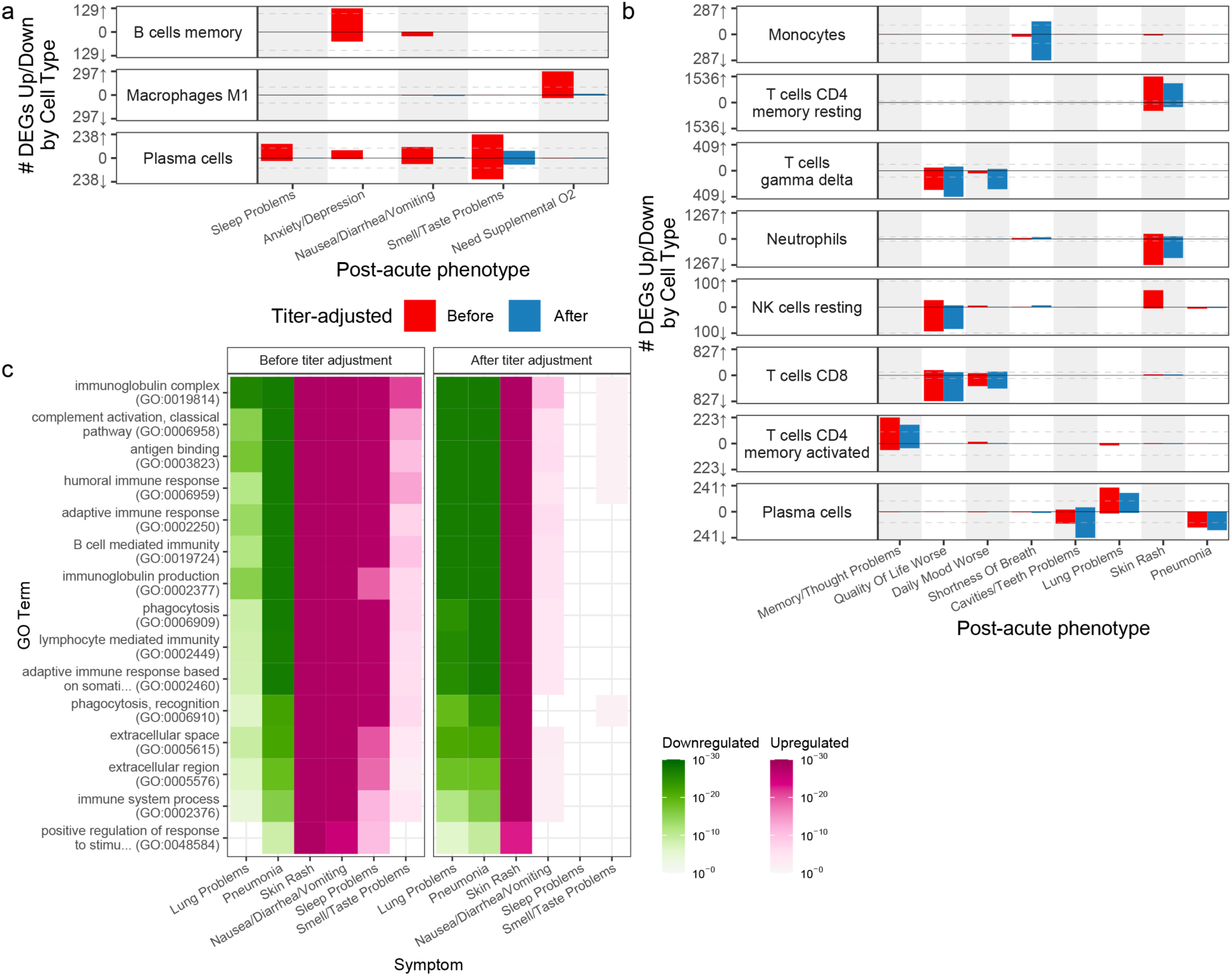
Cell-type-specific differential expression for PASC checklist items. a,b) Anti-spike antibody titer dependent (a) and independent (b) cell-type specific expression signatures. The x axes are PASC checklist items and the y axes the number of upregulated (up arrow) and downregulated (down arrow) DEGs at FDR<0.05. Checklist items are arranged in order of descending prevalence. Each facet presents DE results for the indicated cell type. The dashed grey lines indicate the 100 DEG mark. The color of the bars indicates whether the signatures have been adjusted for anti-spike antibody titers. Only cell types and checklist items with more than 100 dependent/independent DEGs respectively are shown. c) GO term enrichments for plasma cell DEGs. The x and y axes are the checklist items with more than 100 DEGs and GO terms respectively. The union of the top three GO terms for all selected checklist items are shown. The color indicates the direction of the DEGs enriched for that term. Shading of color is representative of the FDR and only FDR < 0.05 are colored. The facets represent before (left) and after (right) controlling for anti-spike antibody titers.

### Patterns of shared DEG signatures

We next examined how DE signatures were shared between pairs of cell-type-specific models (Fig. 4 & Extended Data Fig. 5). Opposite-direction DEGs are defined as the genes upregulated in one model and downregulated in the other, and same-direction DEGs as the genes consistently upregulated or downregulated in both models. In plasma cells, comparing symptoms pairwise for the number of same-direction DEGs revealed two symptom clusters (Fig. 4). Lung problems and pneumonia formed one (“pulmonary cluster”), while sleep problems, nausea/diarrhea/vomiting, skin rash, and smell/taste problems formed the other (“miscellaneous cluster”). Immunoglobulin-related GO terms were downregulated in the pulmonary cluster (Fig. 3c). In the miscellaneous cluster, many of these same terms were upregulated (Fig. 3c). Comparing symptoms between clusters, no same-direction DEGs and significant enrichment for opposite-direction DEGs were observed (Fig. 4), consistent with the infrequent clinical co-occurrence of symptoms in different clusters (Fig. 2c). For quality of life, CD8^+^ and γδ T cells were significantly enriched for same-direction DEGs (Extended Data Fig. 5). Memory resting CD4^+^ T cells and neutrophils had a significant number of opposite-direction DEGs for cavities/teeth problems (Extended Data Fig. 5). Both CD4^+^ T cell and neutrophil DEGs were enriched for CD4^+^ T cell marker genes (CD4^+^ T cell DEG enrichment: odds ratio, OR = 3.5 p = 2.2 × 10e-22; neutrophil DEG enrichment: OR = 2.5, p = 1.04 × 10e-08) and not for neutrophil marker genes (p-values > 0.05), suggesting the neutrophil-specific interaction model is identifying CD4^+^ T cell DEGs. This is consistent with our models capturing opposite-direction DEGs due to the negative correlation between the estimated fractions of these two cell-types (Pearson correlation = -0.60, p = 3.12 × 10e-137).

**Figure 4:**
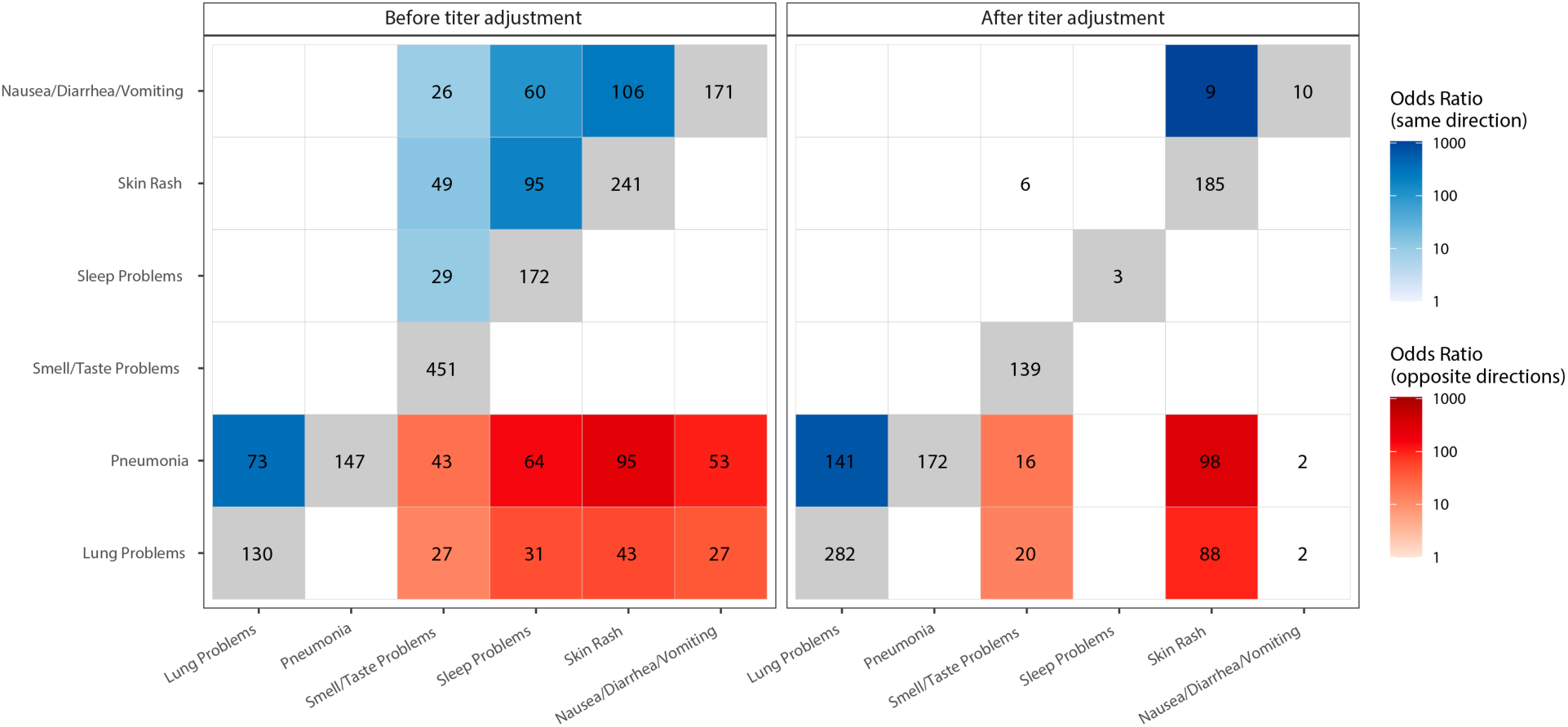
Shared plasma cell DEGs between PASC checklist items. The x and y axes are the PASC checklist items associated with more than 100 DEGs. The numbers in each box are the numbers of shared DEGs between the two checklist items defined in the axes, and the color represents whether they are same-direction (blue), opposite direction (red) or the total number of DEGs for that checklist item (grey). The shadings of red and blue are the odds ratios (ORs) of the Fisher’s exact tests for the enrichment of overlapping genes in that box, and are shown only if the associated enrichment adjusted p-value < 0.05 (FWER, Holm 1979). The left and right facets represent the shared DEGs before and after adjustment for anti-spike antibody titers respectively. Symptoms in rows and columns are ordered by hierarchical clustering and optimal leaf ordering based on the shared same-direction DEGs.

### DEG dependence on anti-spike antibody

Given the DE signatures in plasma cells, whose primary function is to produce antibodies, we assessed whether cell-type specific DEGs were dependent on the antibody response to the SARS-CoV-2 spike protein. To do this, all analyses were repeated, including DE and the verification of our models, while controlling for blood sample titers of anti-spike IgG, IgA, and IgM (Fig. 3, blue bars, Extended Data Fig. 3, Supplementary Table 5). This resulted in a near complete attenuation of both the magnitude and significance of the plasma cell DEG signal for a subset of the miscellaneous cluster (sleep problems, nausea/diarrhea/vomiting, and smell/taste problems), thereby demonstrating a direct link between this subset of PASC and the host-response to SAR-CoV-2 (Fig 3, Extended Data Fig. 6 a-c). For two symptoms (nausea/diarrhea/vomiting and sleep problems), the upregulation of immunoglobulin-related GO terms was no longer observed when controlling for antibody titers (Fig. 3c), consistent with the observed association of titers with sleep problems (Supplementary Table 1). In contrast, lung problems, skin rash, and pneumonia showed little to no attenuation of the plasma cell signal and similar GO term enrichments (Fig. 5b, 3c, Extended Data Fig. 6 d-f). Two additional associations were antibody-dependent: memory B cells with anxiety/depression and M1 macrophages with the need for supplemental oxygen (Fig. 3, Extended Data Fig. 6 g, h). As for DEGs described above, DEGs identified after controlling for anti-spike antibody titers were mostly enriched for the corresponding cell type marker genes from the literature (Supplementary Table 2). The patterns of shared DEG signatures across cell types and symptoms were re-computed after controlling for anti-spike antibody titers. Same-direction DEGs were generally conserved among anti-spike antibody-independent signatures. Notably, the miscellaneous cluster was divided into two components: one entirely antibody-dependent (sleep problems, nausea/diarrhea/vomiting) and one only partially dependent (skin rash and smell/taste problems). The latter subcluster, whose shared DEGs were almost entirely titer dependent, contained many titer independent DEGs (Fig. 4a). Further, both of these symptoms retained their opposite-direction DEGs with the symptoms in the anti-spike independent pulmonary cluster.

## Conclusion

This report presents a transcriptome-wide investigation showing that processes leading to PASC have already started during hospitalization for acute COVID-19. At least two divergent etiologies for different sets of symptoms were identified, one dependent and one independent from the antibody response to the SARS-CoV-2 spike protein. These two points, taken together, suggest that study designs capturing only the post-acute phase risk missing a critical window into the pathogenesis of PASC. The association between acute-phase DE signatures and PASC introduces the potential for discovery of predictive molecular biomarkers. Controlling for the clinical presentation of COVID-19 demonstrates that the molecular processes leading to PASC are not explained simply by acute severity. This is consistent with the reported occurrence of PASC across the range of severity for SARS-CoV-2 infection^1,2^. Additional studies will be required to determine if our findings generalize to mild COVID-19 and asymptomatic infections. It is also anticipated that future studies of the relationship between acute infection and PASC will define additional symptom clusters with common underlying mechanisms. Knowledge of symptom-specific mechanisms will present opportunities to investigate precision treatment and prevention strategies.

Plasma cells are identified as playing a key role in PASC. The downregulation of genes involved in antibody production and function in the pulmonary symptom cluster was independent of the anti-spike antibody titers. The latter are representative of the specific immune response to SARS-CoV-2, thus suggesting a non-specific downregulation of humoral activity underlying pulmonary symptoms. Conversely, the upregulation of genes involved in the same processes in the miscellaneous symptom cluster is largely dependent on the anti-spike antibody response, directly linking the symptoms in this cluster to the immune response to the virus. This antibody dependency could be explained by cross-reactivity of antibodies against SARS-CoV-2^12^. Interestingly, two symptoms in this cluster (skin rash and smell/taste problems) also had separate titer-independent DE signatures, suggesting additional divergent etiologies for these symptoms. Beyond plasma cells, associations with PASC symptoms were seen for CD8^+^ and memory CD4^+^ T cells. The complex pattern of associations seen between cell-type DEGs and symptoms support the hypothesis that PASC is a complex set of traits with multiple etiologies.

## Data Availability

All consented data used will be deposited in public repositories upon publication.

## Acknowledgments

This manuscript is dedicated to study participants of the Mount Sinai COVID-19 Biobank and the healthcare workers who saved their lives. The Mount Sinai COVID-19 Biobank and the work performed here was supported by a redeployed workforce at the Icahn School of Medicine at Mount Sinai, supported by the following centers, programs, departments and institutes: Mount Sinai COVID-19 Informatics Center; Department of Genetics and Genomic Sciences; Human Immune Monitoring Center; Program for the Protection of Human Subjects; Department of Psychiatry; Department of Medicine; Department of Oncological Sciences; Department of Pediatrics; The Precision Immunology Institute; Tisch Cancer Institute; Icahn Institute for Data Science and Genomic Technology; Friedman Brain Institute; Charles Bronfman Institute of Personalized Medicine; Hasso Plattner Institute for Digital Health; Mindich Child Health and Development Institute; Black Family Stem Cell Institute. Natalia Herrera and Steven C. Almo from the Albert Einstein College of Medicine provided the spike protein used in the ELISA measuring the anti-spike antibody titers. S.G., E.G.K, D.M.D.V. and M.M. were supported by NCI U24 grant CA224319. S.G. is additionally supported by grant U01 DK124165. This work was supported in part through the computational resources and staff expertise provided by Scientific Computing at the Icahn School of Medicine at Mount Sinai.

## Author Contributions

The following authors contributed to the overall study design: N.D.B., A.W.C.. The following authors performed all analyses and wrote the manuscript: N.D.B., A.W.C., R.C.T.. The following authors contributed to establishing the Mount Sinai COVID-19 Biobank infrastructure: N.D.B., A.W.C., R.C.T., N.W.S., L.W., E.C., D.M.D.V., G.E.H., B.F., K.M., L.L., J.L.B., C.C., K.N., N.Z., M.A.C., P.R., E.G.K., T.U.M., The Mount Sinai COVID-Biobank Team, S.K.S., M.M., S.G.,E.E.S.. The following authors contributed to the experimental design and procedure for RNA-sequencing: N.D.B., A.W.S., E.C., G.E.H., N.J.F., J.S.J., L.H., T.U.M., S.K.S., R.S., M.M., S.G.,E.E.S.. The following authors contributed to the experimental design and procedure for the anti-spike antibody titers measurement: D.M.D.V., E.G.K., S.K.S., M.M., S.G.. The following authors contributed to data processing and analyses: N.D.B., A.W.C., R.C.T., G.E.H., Y.W., E.G.K., S.G., E.E.S.. The following authors contributed to the mining of electronic medical records for clinical variables: N.D.B., A.W.C., R.C.T., N.W.S., L.W., E.C., D.M.D.V., K.M., M.A.C., B.S.G., G.N., E.E.S.. The following authors contributed to writing specific parts of the text and preparing figures and tables for this manuscript: N.W.S., L.W., E.C., D.M.D.V., A.G.B., B.S.G., E.G.K., S.K.S., R.S., S.G., E.E.S..

## Supplementary Information

Supplementary information is available for this paper.

## Data availability

All consented data used will be deposited in public repositories upon publication.

## Code availability

A modified version of NGSCheckMate suitable for large data sets is available at https://github.com/DarwinAwardWinner/NGSCheckMate. No other custom code was used.

### The Mount Sinai COVID-19 Biobank Team

Charuta Agashe^1^, Priyal Agrawal^1^, Alara Akyatan^1^, Kasey Alesso-Carra^1^, Eziwoma Alibo^1^, Kelvin Alvarez^1^, Angelo Amabile^1^, Carmen Argmann^1^, Kimberly Argueta^1^, Steven Ascolillo^1^, Rasheed Bailey^1^, Craig Batchelor^1^, Priya Begani^1^, Dusan Bogunovic^1^, Swaroop Bose^1^, Cansu Cimen Bozkus^1^, Paloma Bravo^1^, Stacey-Ann Brown^1^, Mark Buckup^1^, Larissa Burka^1^, Sharlene Calorossi^1^, Lena Cambron^1^, Guillermo Carbonell^1^, Gina Carrara^1^, Serena Chang^1^, Steven T. Chen^1^, Jonathan Chien^1^, Mashkura Chowdhury^1^, Jonathan Chung^1^, Phillip H. Comella^1^, Dana Cosgrove^1^, Francesca Cossarini^1^, Liam Cotter^1^, Arpit Dave^1^, Travis Dawson^1^, Bheesham Dayal^1^, Maxime Dhainaut^1^, Rebecca Dornfeld^1^, Katie Dul^1^, Melody Eaton^1^, Nissan Eber^1^, Cordelia Elaiho^1^, Ethan Ellis^1^, Frank Fabris^1^, Jeremiah Faith^1^, Dominique Falci^1^, Susie Feng^1^, Marie Fernandes^1^, Nataly Fishman^1^, Sandeep Gangadharan^1^, Daniel Geanon^1^, Bruce D. Gelb^1^, Joanna Grabowska^1^, Gavin Gyimesi^1^, Maha Hamdani^1^, Diana Handler^1^, Jocelyn Harris^1^, Matthew Hartnett^1^, Sandra Hatem^1^, Manon Herbinet^1^, Elva Herrera^1^, Arielle Hochman^1^, Jaime Hook^1^, Laila Horta^1^, Etienne Humblin^1^, Suraj Jaladanki^1^, Hajra Jamal^1^, Daniel Jordan^1^, Gurpawan Kang^1^, Neha Karekar^1^, Subha Karim^1^, Geoffrey Kelly^1^, Jong Kim^1^, Arvind Kumar^1^, Jose Lacunza^1^, Alona Lansky^1^, Dannielle Lebovitch^1^, Brian Lee^1^, Grace Lee^1^, Gyu Ho Lee^1^, Jacky Lee^1^, John Leech^1^, Michael B. Leventhal^1^, Lora E. Liharska^1^, Katherine Lindblad^1^, Alexandra Livanos^1^, Rosalie Machado^1^, Kent Madrid^1^, Zafar Mahmood^1^, Kelcey Mar^1^, Glenn Martin^1^, Robert Marvin^1^, Shrisha Maskey^1^, Paul Matthews^1^, Katherine Meckel^1^, Saurabh Mehandru^1^, Cynthia Mercedes^1^, Elyze Merzier^1^, Dara Meyer^1^, Gurkan Mollaoglu^1^, Sarah Morris^1^, Emily Moya^1^, Marjorie Nisenholtz^1^, George Ofori-Amanfo^1^, Kenan Onel^1^, Merouane Ounadjela^1^, Manishkumar Patel^1^, Vishwendra Patel^1^, Cassandra Pruitt^1^, Adeeb Rahman^1^, Shivani Rathi^1^, Jamie Redes^1^, Ivan Reyes-Torres^1^, Alcina Rodrigues^1^, Alfonso Rodriguez^1^, Vladimir Roudko^1^, Evelyn Ruiz^1^, Pearl Scalzo^1^, Ieisha Scott^1^, Sandra Serrano^1^, Hardik Shah^1^, Mark Shervey^1^, Pedro Silva^1^, Laura Sloofman^1^, Melissa Smith^1^, Alessandra Soares Schanoski^1^, Juan Soto^1^, Shwetha Hara Sridhar^1^, Hiyab Stefanos^1^, Meghan Straw^1^, Robert Sweeney^1^, Alexandra Tabachnikova^1^, Collin Teague^1^, Manying Tin^1^, Scott R. Tyler^1^, Bhaskar Upadhyaya^1^, Akhil Vaid^1^, Verena Van Der Heide^1^, Natalie Vaninov^1^, Konstantinos Vlachos^1^, Daniel Wacker^1^, Laura Walker^1^, Hadley Walsh^1^, Bo Wang^1^, Wenhui Wang^1^, C. Matthias Wilk^1^, Jessica Wilson^1^, Karen M. Wilson^1^, Hui Xie^1^, Li Xue^1^, Naa-akomaah Yeboah^1^, Nancy Yi^1^, Mahlet Yishak^1^, Sabina Young^1^, Alex Yu^1^, Nina Zaks^1^, Renyuan Zha^1^

1. Icahn School of Medicine at Mount Sinai, New York, NY, 10029, USA

## Methods

### Ethics Statement

This study was approved by the Human Research Protection Program at the Icahn School of Medicine at Mount Sinai (STUDY-20-00341). All patients admitted to the Mount Sinai Health System were made aware of the research study by a notice included in their hospital intake packet. The notice outlined details of the specimen collection and planned research. Flyers announcing the study were also posted in the hospital and a video was run on the in-room hospital video channel. Given the monumental hurdles of consenting sick and infectious patients in isolation rooms, the Human Research Protection Program allowed for sample collection, which occurred at the time of clinical collection and included at most an extra 5-10 cc of blood, prior to obtaining research consent. Limited existing clinical data obtained from the medical record was collected and associated with the samples. As the research laboratory processing needed to begin proximal to sample collection, a portion of the data was generated prior to obtaining informed consent. During or after hospitalization, research participants and/or their legally authorized representative provided consent to the research study, including genetic profiling for research and data sharing on an individual level. In those circumstances where consent could not be obtained (13.8% of subjects, 0% of subjects who completed the post-discharge checklist), data already generated could continue to be used for analysis purposes only when not doing so would have compromised the scientific integrity of the work. The data were not identifiable to the researchers doing the analyses.

### Sample Collection

Patients presenting to the Mount Sinai Health System (MSHS) between April and June of 2020 were enrolled through daily manual review of new hospitalizations for COVID-19. Blood collection was performed in conjunction with routine clinical blood draws throughout the participants’ hospital stay. Research specimens were brought to biosafety level 2 plus facilities for accessioning, processing, and storage of serum, plasma, whole-blood, and peripheral blood mononuclear cells (PBMCs). The whole blood utilized for RNA sequencing (RNA-seq) was collected in Tempus RNA Blood Tubes (ThermoFisher, #4342792). As soon as possible after blood collection, tubes were shaken and stored at -80ºC. Blood used for Olink and enzyme-linked immunosorbent assays (ELISA) were collected in SST tubes (Becton Dickinson #367985) and blood used for whole genome sequencing (WGS) in CPT Vacutainer tubes (Becton Dickinson #362761). Blood in SST tubes was centrifuged to extract serum that was then aliquoted and stored at -80ºC in Cryovials (Crystalgen #19335-6SPR). Blood from CPT tubes was aliquoted for WGS and stored at -80ºC in Cryovials (Crystalgen #19335-6SPR).

### Enzyme-linked immunosorbent assay

We modified a previously reported protocol for performing ELISA on all blood samples [doi: 10/gj9rth]. Briefly, 96-well plates were coated at 4ºC overnight with 100 μl per well of a 1 mg/mL solution of recombinant spike protein against the RNA binding domain region suspended in phosphate-buffered saline (PBS; Sigma-Aldrich). The coating solution was then removed and wells were washed 3 times with 100 μl of washing buffer (PBS with 0.05% v/v Tween 20). Next, 150 μl of coating buffer (PBS with 1% w/v bovine serum albumin (endotoxin-free)) was added to each well and incubated at room temperature for 1 hour to block wells. During blocking, plasma samples were diluted 1:200 in blocking buffer. Plates were then washed three times with 100 μl per well of wash buffer. Next, a 1:3,000 dilution of goat anti-human IgG/A/M F(ab)– horseradish peroxidase conjugated secondary antibody was prepared in PBS and 100 μl of this secondary antibody was added to each well for 1 hour (SouthernBiotech #2020-04 Lot No. L4206-Q408B, #2050-04 Lot No. C5213-RI66P, #2040-04 Lot No. B3919-NE80C). Plates were washed three times with a wash buffer. Once dry, 100 μl SIGMAFAST OPD (o-phenylenediamine dihydrochloride; Sigma–Aldrich) solution was added to each well. This substrate was left on the plates for 5 minutes and reaction was stopped by the addition of 50 μl per well of 3 M hydrochloric acid. The optical density at 580 nm (OD450) was measured using a Synergy 4 (BioTek) plate reader.

### Olink data generation

Serum samples were analyzed for a panel of 92 circulating proteins associated with human inflammatory conditions using the Olink multiplex assay (Olink Target 96 Inflammation, Olink Bioscience, Uppsala, Sweden) according to the manufacturer’s instructions. Incubation master mix containing pairs of oligonucleotide-labeled antibodies to each protein was added to the samples and incubated for 16 hours at 4ºC. Each protein was targeted with two different epitope-specific antibodies to increase the specificity of the assay. Presence of the target protein in samples brings partner probes in close proximity to each other, allowing formation of a double-stranded oligonucleotide polymerase chain reaction (PCR) target. The following day, extension master mix was added to samples to cause specific target sequences to be detected and generate amplicons using PCR in a 96 well plate. For the detection of specific protein, a Dynamic Array integrated fluidic Circuit 96×96 chip was primed, loaded with 92 protein-specific primers, and mixed with sample amplicons including three inter-plate controls and three negative controls. Real-time microfluidic quantitative PCR was performed in Biomark (Fluidigm, San Francisco, CA) for target protein quantification. Data was analyzed using real-time PCR analysis software via the ΔΔCt method [doi:10/c689hx] and NPX (Normalized Protein Expression) manager. Data were normalized using internal controls in each sample, inter-plate controls to normalize across plates, and a correction factor calculated by Olink from negative controls, producing NPX values proportional to the log2 of the protein concentration^13^.

### Clinical Data Generation

Clinical data elements (CDE) analyzed for this report were ascertained by employing a three-pronged strategy. First, automated extraction of structured CDE from electronic health records (EHR) was employed, constructing an exhaustive database of over one thousand CDE that included demographics, vitals, comorbidities, clinical laboratory test results, and medications. For CDEs with multiple entries in a 24-hour window, entries were collapsed by calculating the median for numerical CDE and retaining the most severe entry for categorical CDE. Groups of CDE were combined to derive more complex CDE, such as 24-hour summaries of COVID-19 severity (defined as 4 categories: control (i.e. SARS-CoV-2 negative), moderate, severe, and severe with end organ damage (EOD))^14^. Second, we employed manual chart review by subject matter experts to extract unstructured CDE from the free text of clinical notes such as dates of COVID-19 symptoms onset. For these two components of our CDE ascertainment strategy, manual and automated quality control checks were performed to verify consistency and correctness of the data. The third approach to CDE ascertainment was a checklist of health changes after COVID-19 (or, for controls, after hospitalization). The checklist was constructed when anecdotal reports of post-acute sequelae were just beginning to surface with the goal of capturing a set of CDE that collectively reflected the prevailing view of PASC at the time (Supplementary Table 1). Checklists were completed by study participants remotely via a webform following the acute COVID-19 hospitalization with a clinical research coordinator on the phone for assistance. The analyses in this report utilized checklists completed from February 2021 through July 2021. Checklist items were generally of two types, those that assessed clinical deterioration (e.g., quality of life worsening since COVID-19, “general”) and those that assessed symptom presence (e.g., the emergence of memory issues since COVID-19, “symptoms”). All were coded as boolean variables, with “true” indicative of either clinical deterioration or the presence of the symptom. We removed symptoms reported by N≤5 subjects with RNA-seq from further analyses to allow models to converge. At the time of the data freeze for this report, a small number of participants had completed the checklist on more than one post-acute timepoint as part of an ongoing effort to chronicle post-acute trajectories. In such instances, answers were collapsed into a single value, with a “true” value for any item coded as such at any of the timepoints. All individuals without any survey answers were assigned to a third “unknown” group for each item. For comparing proportions of reported symptoms between COVID-19 patients and hospitalized controls, we performed a one-sided Fisher test for enrichment, correcting for multiple testing using the Benjamini-Hochberg (BH) method for false discovery rate (FDR) control^15^.

### Selection of RNA sequencing batch controls

Technical effects that emerge from the batching of samples at various processing steps in gene expression studies (e.g., extraction, sequencing) are a large confounding variable in downstream analyses. This is often controlled by constructing batches using a randomization procedure that balances key outcome variables (e.g., case/control status) across batches. Ideally, randomization is performed when the full set of samples to be analyzed has been collected. For the current study, sample collection and sequencing occurred in parallel so as to rapidly generate data to study the host response to SARS-CoV-2 infection. To account for batch effects without masking signal of interest, eight samples were chosen as “batch controls” to be included in every sequencing batch of 192 samples. The sample size of eight batch controls was determined based on simulation analyses that suggested this number would be sufficient to control for batch effects (data not shown). Batch controls were manually selected to be a representative subset of the full cohort of samples with respect to key technical (e.g., RNA quality) and biological (e.g., COVID-19 status) variables.

### Randomization of RNA-seq batches

After selecting batch controls, samples were randomized into batches for RNA-seq (batch size of 192) and extraction (batch size of 8 created within each sequencing batch of 192). One million permutations of batch assignments were performed. The permutation that minimized the mean canonical correlation between batch assignment and the following set of variables was selected: time point, age, ethnicity, race, sex, COVID-19 status, deceased flag, intensive care unit status, ventilation status, intubation status, batch control status, and blood volume collected. Randomization was done in multiple phases during sample collection, each phase performed on the set of samples that had been collected since the previous phase, thereby maximizing the degree of randomization that could be achieved while allowing sequencing to proceed in parallel with sample collection. Batch control samples were part of the randomization but were forced to be present in every sequencing batch.

### RNA Extraction, Library Preparation, and Sequencing

RNA extraction, library preparation, and sequencing were performed as described previously^16^. Briefly, frozen blood samples were thawed and total RNA was extracted from the samples using a modification of the MagMax protocol for Stabilized Blood Tubes RNA Isolation Kit (Thermo Fisher, #4451893). Samples that yielded sufficient RNA (>50 ng) were barcoded and prepared for pooled whole transcriptome sequencing using the TruSeq Stranded Total RNA Library Prep Gold (Illumina, #20020599), which is designed to remove ribosomal, globin, and mitochondrial RNA. Libraries were amplified with 15 cycles of PCR, pooled, and sequenced on a NovaSeq 6000 (Illumina) using Sprime flow cells with 100 base pair paired-end reads, targeting a mean of 50 million read pairs per sample. For a minority of samples in which the first extraction failed (N=24), RNA was re-extracted from the supernatant saved from the first centrifugation pellet. The extraction protocol was repeated starting with the second wash step after re-pelleting the RNA.

### DNA Extraction

After blood collection, tubes were shaken and whole peripheral blood aliquoted into cryovials and stored at -80C. The MagMax DNA Multi-Sample Ultra 2.0 Kit protocol (ThermoFisher, #A36570) was used to isolate DNA from 0.2 mL of collected peripheral blood, following the manufacturer’s instructions. To facilitate DNA extraction, a KingFisher Flex machine was used to automate the isolation of 96 samples at once with the MagMAX_Ultra2_200μL_FLEX program. Briefly, frozen blood cryovials were thawed at room temperature prior to DNA extraction. Next, processing plates were labeled and assembled. Plate 1 contained 500 μL of Wash 1 Solution, Plate 2 contained 500 μL of Wash 2 solution, Plate 3 contained 500 μL of Wash 2 solution, and Plate 4 contained 75 μL of elution buffer. The sample plate was then prepared by first adding 20 μL of enhancer solution, then 200 μL of the peripheral blood sample, and 20 μL of proteinase K in that order. All plates were then put into the KingFisher to start DNA extraction. In the middle of the program, 220 μL of DNA Binding bead mix was added to each sample well. At the end of the run, the elution plate was removed from the instrument and the DNA samples were transferred to a skirted 96-well plate. If there were excess beads in the DNA samples, the beads were collected on a plate magnet and the purified DNA samples were transferred into a new skirted 96-well plate.

### Whole Genome Sequencing

Once the isolated DNA passed quality control, we conducted WGS library preparation with the Nextera DNA Flex Library Preparation Kit (Illumina, #20018705), using 250-500 ng of genomic DNA as input and by following the manufacturer’s protocol. In brief, the genomic DNA samples were simultaneously fragmented and ligated with adapters by tagmentation. The tagmented DNA fragments were then amplified using a limited number of PCR cycles to ligate indexes to each template. The quality of the final libraries was then validated on the Agilent TapeStation 4200 using High Sensitivity D1000 screen tape (Agilent Technologies, #G2991AA). The concentration of each library was measured on a Quant-iT High Sensitivity dsDNA Assay Kit (ThermoFisher Scientific, #Q33120). Following library preparation, we performed WGS targeting 30X coverage using Illumina paired-end, short-read sequencing technology on the NovaSeq 6000 instrumentation. To achieve even coverage across patient genomes, these libraries were sequenced at a multiplex of 24-29 samples per batch and assigned to the S4 NovaSeq flow cell to account for batch size. This configuration enabled 2×150 bp paired end reads into resulting FASTQ files in 2-5 days per batch that were sent through primary data quality control using MultiQC to assess read depth and quality metrics^17^.

### Alignment and Quantification of RNA-seq data

After RNA sequencing data collection, base calls were converted into raw reads and filtered after quality assessment. Quality-filtered raw data was converted into FASTQ files using bcl2fastq (Illumina). RNA-seq reads were aligned to the GRCh38 primary assembly^18^ with Gencode gene annotation v30^19^ by STAR (v2.7.3a)^20^ using per-sample 2-pass mapping (--twopassMode Basic) and chimeric alignment options (--chimOutType Junctions SeparateSAMold -chimSegmentMin 15 -chimJunctionOverhangMin 15). RNA-seq QC metrics were calculated by fastqc (v0.11.8) and Picard Tools (v2.22.3). Quantification was done at the gene-level with antisense specificity using featureCounts (Subread R package v1.6.3 and strandness option -s 2)^21^ with gene-level grouping / primary alignments only / count all overlapped features (-t exon -g gene_id -primary -O). MultiQC^17^ was used to compile and summarize per-sample statistics from STAR, Picard Tools and featureCounts (i.e. gene-level counts, mtRNA counts, globinRNA counts, etc.) into an interactive HTML report.

### Sample Mislabelling Correction

To address sample mislabeling, we assembled several sources of information to enable identification of mislabeled samples and inference of correct labels. For each RNA-seq sample, we defined expressed sex based on the relative abundance of the sex-specific genes UTY (male) and Xist (female). NGSCheckMate was used to determine which samples were empirically derived from the same subject based on correlation between variant allele fractions at a set of pre-specified loci (genetic match)^22^. These data were used to identify discrepancies between label matches and genetic matches and infer the correct subject labels. In cases where correct labels could not be unambiguously inferred from the RNA-seq and WGS, the ambiguity was resolved by identifying samples that showed aberrant patterns in the ELISA and Olink data, such as a single ELISA sample with substantially lower titers than both the samples before and after it from the same subject, or an Olink sample that failed to cluster with other samples from the same subject when visualized in Clustergrammer^23^. In most cases correct labels could be unambiguously inferred, even in complex cases involving multiple overlapping mislabeling events. Any mislabeled samples for which correct labels could not be inferred were discarded from all analyses.

### RNA-seq count data processing

DV200, the percentage of fragments longer than 200 nucleotides, has been shown to be more reliable than RNA integrity number (RIN) to assess quality in RNA-seq data^24^. We therefore excluded samples with DV200 below 80% as well as samples with fewer than 10 million mapped reads counted by featureCounts. Despite globin depletion during library preparation, some samples showed substantial read counts for globin genes. To remove the unwanted signal due to globin gene expression in whole blood, counts for all annotated globin genes (gene symbols CYGB, HBA1, HBA2, HBB, HBD, HBE1, HBG1, HBG2, HBM, HBQ1, HBZ, and MB) were discarded, and the remaining count matrix was transformed to counts per million (CPM). Genes with CPM >=1 in >=36 samples (half the number of subjects with no positive PCR or antibody test for SARS-CoV-2 during the study period) were included in our analyses (21,194). Gene expression was normalized for composition bias using the trimmed mean of M-values method, implemented by calcNormFactors in the edgeR package^25,26^ and transformed to normalized log2 CPM with observation weights computed by voomWithDreamWeights from the variancePartition package^27,28^.

RNA-seq data often contains technical and biological sources of variation irrelevant to the question at hand. Exploration of the variance in the gene expression data was performed with principal component analyses (PCA, prcomp R function) and variance partition analyses^29^. Starting from normalized counts, we identified the variable that was the next strongest driver of unwanted variance, adjusted for this variable using linear modeling along with all previously selected ones, and repeated this procedure iteratively until no more confounding variables were observed to be strong drivers of variance in the data. This resulted in a set of non-redundant technical and biological covariates explaining a substantial fraction of the unwanted variation in the gene expression. To further identify covariates that might have an important impact on the gene expression data in a way that could not be easily captured by these analyses, we leveraged WGCNA co-expression network analyses^30^. We started by fitting a linear mixed model to the log2 CPM values including all the previously selected variables using dream^27^ and extracted the residuals from this model. We then built a co-expression network from the residualized expression values and selected a new variable that was significantly correlated to module eigengenes of a large number of modules after multiple testing correction (Bonferroni-adjusted p-value < 0.05). The data was then residualized again, including the newly selected variable in the model, and the process was repeated until no more confounding variables were observed driving substantial variation in any modules.

Using this approach, we identified a set of technical and biological confounding variables that we used for all analyses performed. Specifically, whenever fitting a linear mixed model, we accounted for the following covariates as fixed effects: number of days since the first blood sample, RNA DV200, Age, PCT_R2_TRANSCRIPT_STRAND_READS, PCT_INTRONIC_BASES, WIDTH_OF_95_PERCENT; as well as accounting for the following covariates as random effects: Subject_ID and Sex. The number of days since the first blood sample was modeled as a smooth nonlinear function defined using natural cubic splines (ns R function^31^) with internal knots at 1, 3, 7 and 12, reflecting the timeline of blood draws for individuals in our cohort. All other fixed effect technical variables were scaled to have a mean of 0 and a variance of 1 using the scale R function. The last 3 fixed effects listed are sequencing quality metrics computed by Picard Tools: PCT_R2_TRANSCRIPT_STRAND_READS is “the fraction of reads that support the model where R2 is on the strand of transcription and R1 is on the opposite strand”; PCT_INTRONIC_BASES is the “fraction of PF_ALIGNED_BASES that correspond to gene introns”; and WIDTH_OF_95_PERCENT is difference between the 2.5 percentile and the 97.5 percentile of the insert size distribution.

PCA was performed on the residualized expression matrix after adjusting for all covariates selected above using linear mixed model for all the samples, and outliers were removed by drawing an ellipse in the first 2 PCs with semi-major and semi-minor axes equal to 3 standard deviations in PCs 1 and 2 respectively, then discarding all samples outside this ellipse. Batch effects were residualized from the data by fitting a linear mixed model to the normalized log2 CPM and weights for each gene with random effects for library prep plate (the batch effect to be residualized out) and blood sample ID (the biological signal to be retained) using dream^27^, and then subtracting the fitted effects for library prep plate while retaining the fitted effects for blood sample and the residuals. Then, the technical replicates for each batch control sample were summarized to a single residualized expression value equal to the weighted mean of all technical replicates, and a single weight equal to the sum of the weights of all technical replicates. This yielded a batch-residualized expression matrix with 21,194 rows (genes) and 1,392 columns (blood samples) and a corresponding matrix of observation weights that we used as the input for all differential expression testing.

### Cell Type Deconvolution and Validation

Cell type fractions were estimated for each sample using CIBERSORTx^32^, providing transcripts per million (TPM)^33^ as input and following procedures recommended by the documentation. For batch-controls, reads from all technical replicates were pooled before computing TPM. CIBERSORTx requires a reference data set, which it uses to determine the set of possible cell types into which expression values may be deconvolved as well as the set of cell-type specific genes that will be used for deconvolution. To ensure the most accurate estimation, we tested 4 independent references generated by different labs with different technologies. The LM22 reference consists of bulk RNA-seq data from PBMCs sorted by fluorescence-activated cell-sorting, while the NSCLC PBMC, SCP424, and Wilk references are derived from single-cell RNA-seq of PBMCs in various disease contexts^34-36^. The SCP424 data set was pre-processed as previously described^16^. The Wilk reference consisted of a list of genes upregulated in 20 different cell types (relative to the other 19), and genes with adjusted p-value below 0.05 for a given cell type were defined as markers for that cell type in our analyses.

For validation purposes, the cell type fractions estimated with each reference were grouped into neutrophils, monocytes, and lymphocytes. Then the sum of each group was compared against the corresponding complete blood count fraction recorded on the day of sample collection using Pearson correlation. The cell types for LM22, NSCLC PBMC, and SCP424 were grouped as described previously^16^, while the groupings for the Wilk reference were as follows: for monocytes, “CD14 Monocyte” and “CD16 Monocyte”; for lymphocytes, “CD8m T”, “CD4m T”, “B”, “IFN-stim CD4 T”, “Proliferative Lymphocytes”, “γδ”, “IgM PB”, “IgG PB”, and “IgA PB”; for neutrophils, just “Neutrophil”. The LM22 reference cell type fractions had consistently the highest correlation and were selected for all further analyses.

### Cell type selection

To control for variations in cell type fractions between samples, we identified a minimal set of cell type fractions explaining the variation in severity, to include as covariates when fitting models for differential expression. To account effectively for the biggest driver of variance in the cell type fractions in our data, Subject ID, and all other identified confounders, we used glmmLasso to fit an L1-penalized ordinal regression generalized linear mixed model to all estimated cell type fractions, with severity as the response using the adjacent categories family (glmmLasso function with options family = acat(), final.re = TRUE, and switch.NR = TRUE)^37^. We optimized the tuning parameter lambda across a range from 0 to 500 counting by 5 and found that Bayesian information criterion^38^ was minimized at lambda = 95. Finally, we determined a set of non-redundant cell types explaining severity by selecting all cell types with non-zero parameters after penalization.

To select which cell types would be tested in the cell fraction interaction model described below, we used variancePartition to determine the contribution of COVID-19 severity to the variation observed in each cell type. We ran fitVarPartModel and extractVarPart on the cell type fractions with a model including all identified confounders as well as severity as a random effect to calculate the fraction of variance explained in each cell type by each term in the model^29^. Cell types in which severity explained at least 1% of the variance and which had non-zero fractions in at least 20% of samples were selected for differential expression testing with the interaction model.

### Differential Expression Analyses

For each PASC checklist item, we fit a linear mixed model to the batch-corrected expression of each gene, controlling for all previously identified confounders, COVID-19 severity at the time of sampling and any ICU encounter during their hospital stay as random effects and selected cell type fractions as fixed effects. We tested each gene for differences in expression between the “true” and “false” groups using dream^27^. In addition, for each combination of checklist item and cell type, we fit the same model with an additional fixed effect interaction term between the checklist item and the cell type fraction. This interaction model estimates, for each group of the checklist item, a coefficient for the slope of gene expression with respect to the specified cell type fraction in that group. We tested for differences in these slope coefficients for the “true” and “false” groups, therefore looking for genes whose expression are varying with the cell type fraction in different ways between the groups. Lastly, we tested for differences in gene expression that are independent of the antibody response to the spike protein by fitting all models a second time with 3 additional coefficients controlling for the log2 titers of anti-spike-protein IgG, IgA, and IgM. For this, we included all 1301 samples from 543 subjects that had both RNA-seq and serology measures (403 samples from 176 subjects with PASC checklists). For each differential expression test, we controlled for multiple testing among the 21,194 genes tested using the BH method15. We only describe in the main text cell-type/symptom combinations that have at least 100 differentially expressed genes at FDR ≤ 0.05.

### Shared DEG analysis

We defined the same-direction shared DEGs between two DE tests to be the set of genes that are differentially expressed in both tests and have the same sign on their logFC values (i.e. both negative or both positive). Similarly, we defined the opposite-direction shared DEGs as the set of genes differentially expressed in both tests but with opposing signs (i.e. negative in one test and positive in the other). We tested for enrichment in shared DEGs by performing a one-sided Fisher’s exact test for enrichment of either the same-direction or opposite-direction shared DEGs relative to all the DEGs for each of the two DE tests. We controlled for multiple testing using Holm’s method for family-wise error rate control^39^ among all comparisons performed within a given symptom or cell type between DE signatures with at least 100 DEGs either before or after controlling for anti-spike antibody titers.

### Gene Ontology term enrichment analyses for DE signatures

For each DE test, the downregulated and upregulated DEGs were separately tested for Gene Ontology (GO) term enrichment for all GO terms annotated to at least 10 expressed genes, using the Bioconductor packages goseq, topGO, and org.Hs.eg.db. A one-sided Fisher’s exact test was performed for the enrichment of the up- or downregulated DEGs for each GO term, with all 21,194 expressed genes as the background. For each enrichment analysis, we controlled for multiple testing among all GO terms tested using the BH method^15^.

### Cell type specific marker gene enrichments

To verify that the interaction models between PASC checklist items and cell-type fractions captured cell-type-specific DEGs, we tested for enrichment of cell-type-specific marker genes. For each of several broad cell-type categories (Supplementary Table 6), we assembled a set of marker genes from the literature as the union of all marker genes for each category^36,40-42^. We defined the list of DEGs for each category as the union of the DEGs from all PASC symptoms for all LM22 cell types in that category and tested whether this union of DEGs was enriched for the marker genes using a one-sided Fisher’s exact test. P values were adjusted for multiple testing using the BH method^15^.

### Serology, acute-phase CDE, and cell fraction dream models

For each estimated cell-type fraction, we tested for differences between the “true” and “false” groups of each checklist item using dream in the same manner as for the gene expression data, except that coefficients for the cell-type fractions themselves were omitted from the model. Likewise, we tested for differences in each CDE measured during hospitalization, omitting the coefficients for the cell-type fractions and all RNA-related coefficients (RNA DV200, PCT_R2_TRANSCRIPT_STRAND_READS, PCT_INTRONIC_BASES, and WIDTH_OF_95_PERCENT). Lastly, we tested for differences in log2 titers of anti-spike IgG, IgA, and IgM using the same model as for the CDE.

### Other data processing, analyses, and visualization

Most data analyses were performed using the R statistical language version 4^43^ and Bioconductor suite of packages^44^. Large data tables were read, written, and processed using many tidyverse packages^45^ as well as the R package data.table. Multiple testing correction was performed using the p.adjust R function. Unless otherwise noted, all plots were prepared using ggplot2^46^. Fisher’s exact tests were performed using the fisher.test R function. Correlation tests were performed using the cor.test R function. Analyses were run in parallel using the R packages future, BiocParallel, foreach, doMC, batchtools^47^, and parallelDist. Intermediate results were cached for faster re-analysis using the R packages memoise and cachem. Study data were collected and managed using REDCap electronic data capture tools hosted by Scientific Computing at the Icahn School of Medicine at Mount Sinai^48,49^. Rows and columns in correlation plots and shared DEG plots were ordered using the R package seriation^50-53^. GO term enrichment results were clustered with GO-Figure!^54^ and visualized using the R package treemap. Canonical correlations between all technical, clinical, and demographic variables were calculated using the canCorPairs function and visualized using the plotCorrMatrix function from the Bioconductor package variancePartition^29,55^.

## Extended Data Figures

**Extended Data Figure 1:**
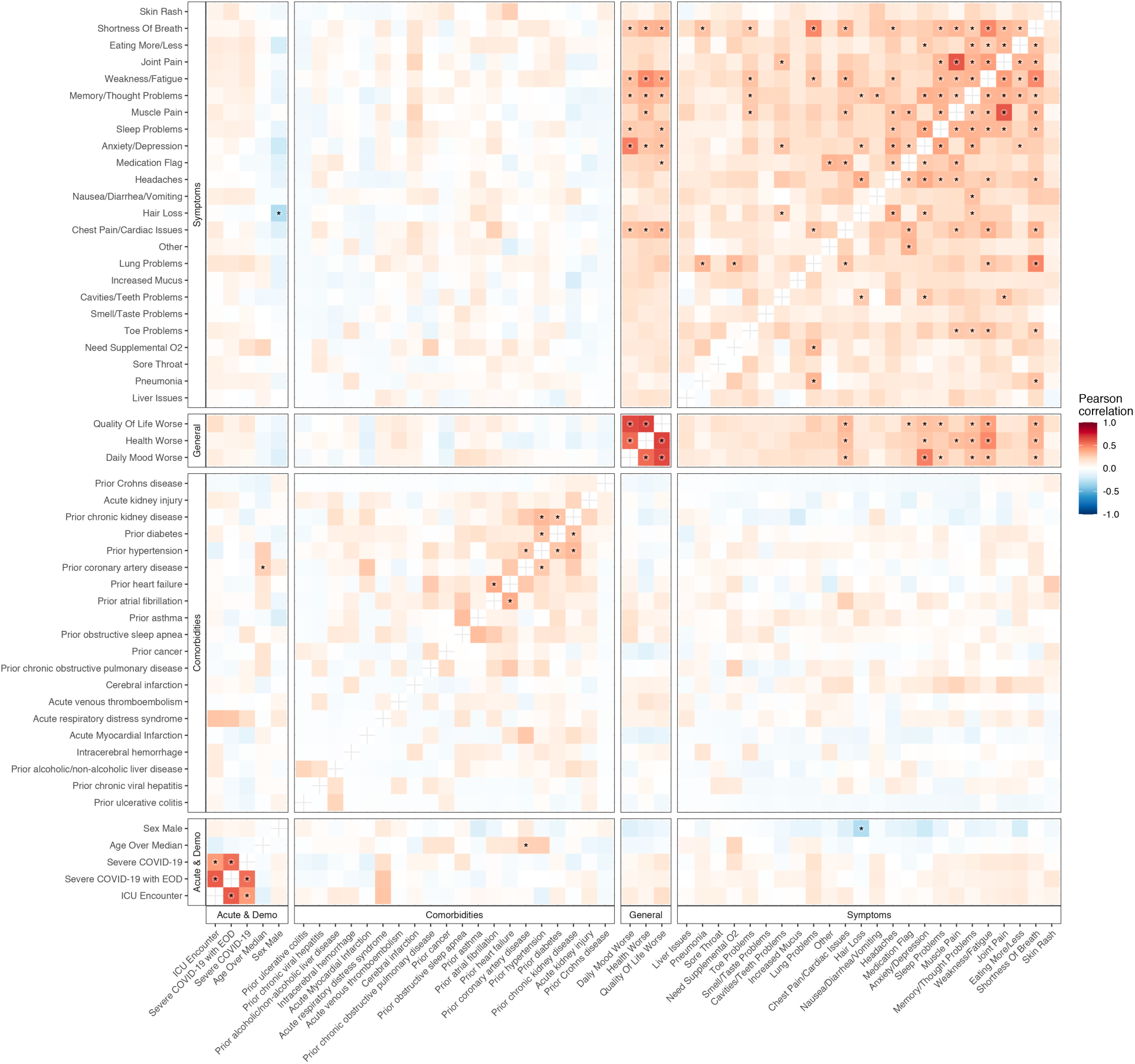
Correlation of occurrences of PASC checklist items, comorbidities, demographics, and acute disease metrics. The axes are representative of the symptoms, comorbidities, demographics, and acute disease metrics, and the color represents the Pearson correlation of their coincidence. Comorbidities present before COVID-19 hospitalization are defined with the prefix “prior” in the axis label. Correlations with family wise error rate (FWER, Holm 1979) adjusted p values < 0.05 are indicated with a star. Rows and columns are ordered by hierarchical clustering and optimal leaf ordering.

**Extended Data Figure 2:**
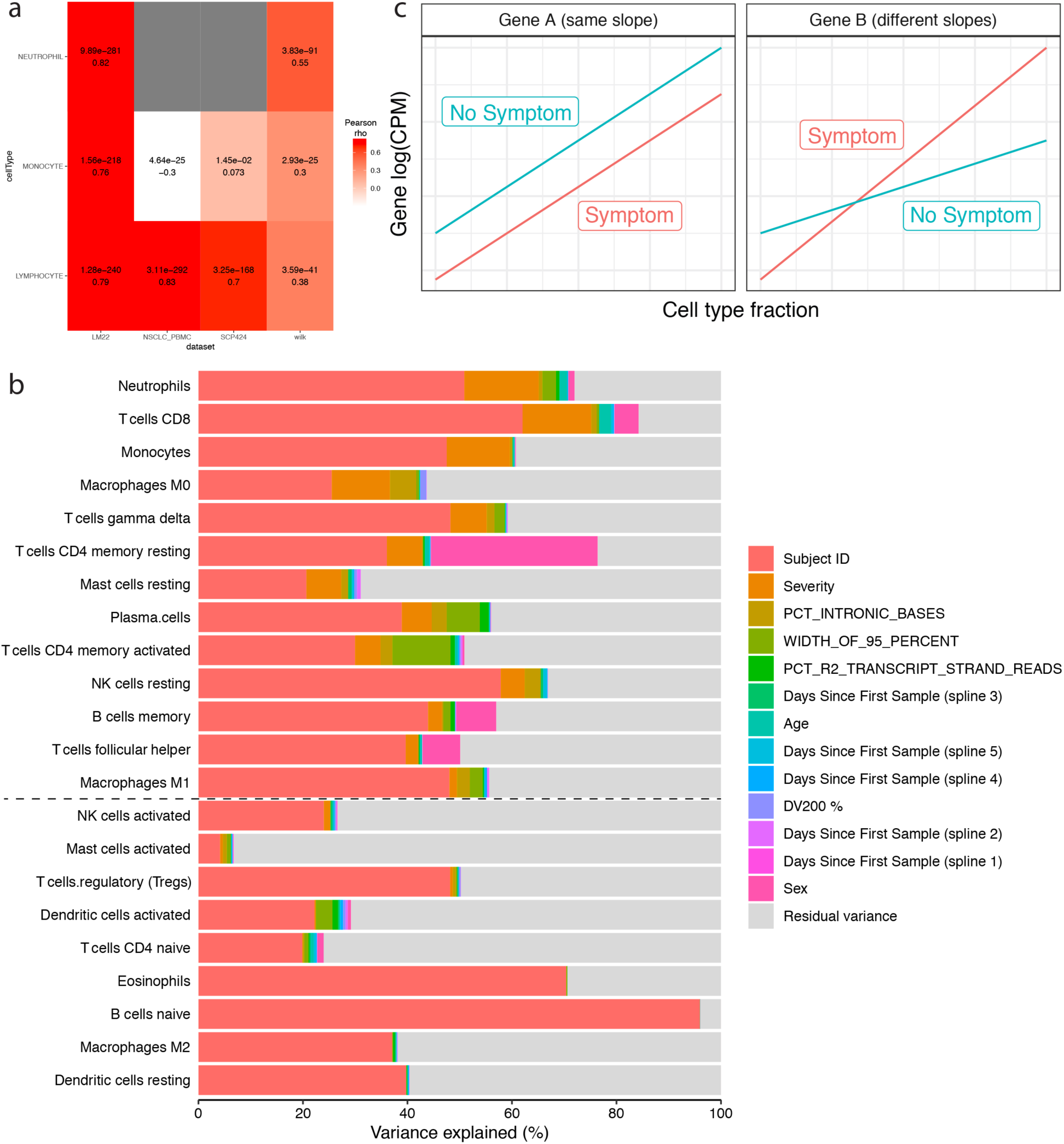
Cell-type fraction estimations and interaction model. a) Validation heatmap of estimated cell-type fractions with clinical complete blood counts. The x axis shows the literature reference dataset used for the deconvolution procedure and the y axis is the cell-type fractions validated. The colors represent the Pearson correlation values between the estimated cell-type fractions and the corresponding complete blood count from the clinical data. The adjusted p-values (FWER, Holm 1979) and correlation values are noted in each box. b)Estimated cell-type fraction variance explained by biological and technical variables. The x axis is the percent of variance of the cell-type fractions explained by covariates (colors) and the y axis the cell type assessed. Cell types are ordered by the decreasing percent of their variance explained by COVID-19 severity. The black dashed line represents the cutoff for inclusion in the cell-type-specific analyses. c) Schematic of interaction model for mock genes A and B. The x axis is the cell-type fraction of a specific cell-type of interest and the y axis the gene expression in log2(counts per million). The color represents the presence (red) and absence (blue) of a symptom. The left and right facets show a gene not differentially expressed (same slope) and a differentially expressed gene (different slopes) respectively.

**Extended Data Figure 3:**
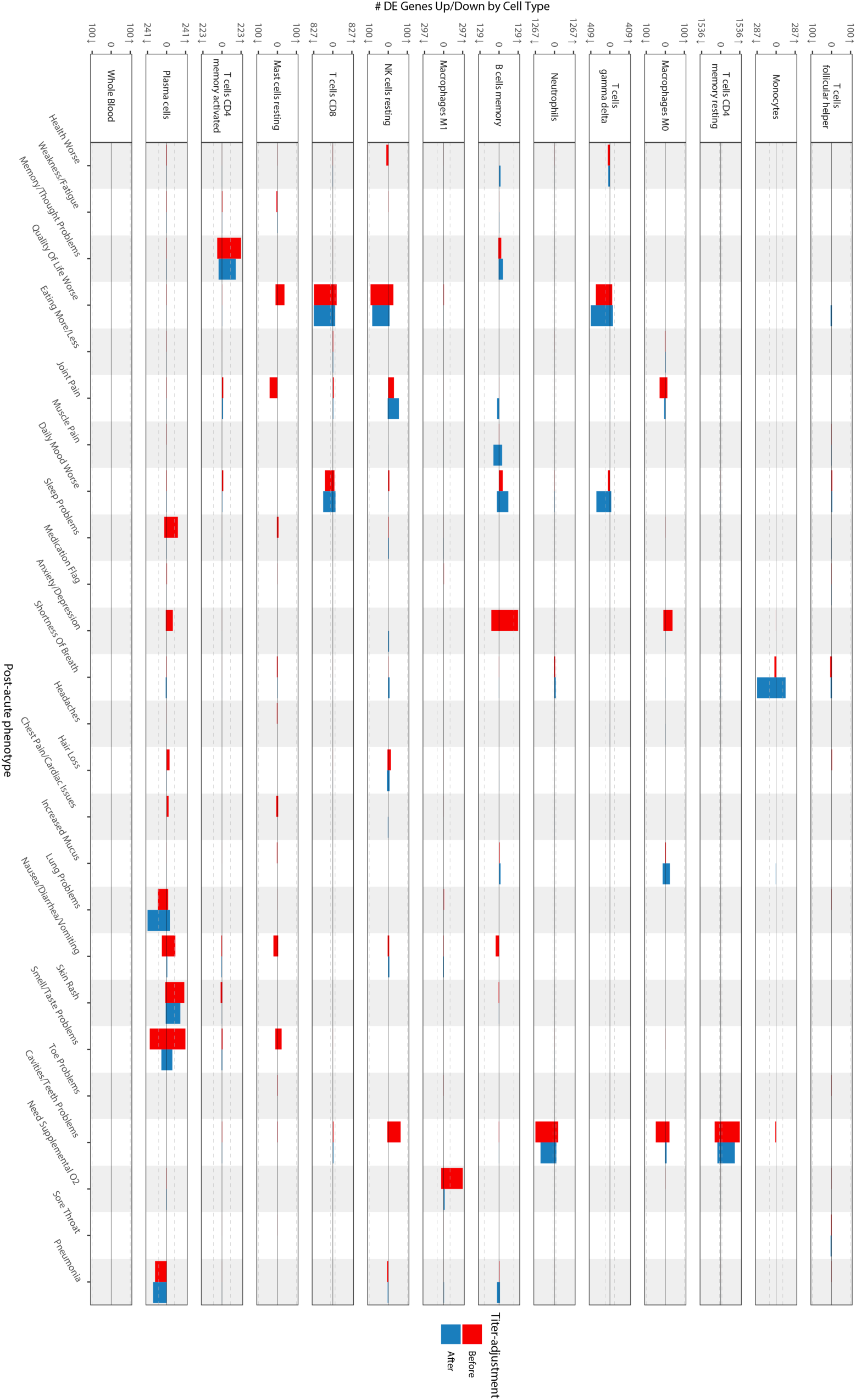
Cell-type-specific differential expression for PASC checklist items (full) The x axes are PASC checklist items and the y axes the number of upregulated (up arrow) and downregulated (down arrow) DEGs at FDR<0.05. Checklist items are arranged in order of descending prevalence. Each facet presents DE results for the indicated cell type. The dashed grey lines indicate the 100 DEG mark. The color of the bars indicates whether the signatures have been adjusted for anti-spike antibody titers.

**Extended Data Figure 4:**
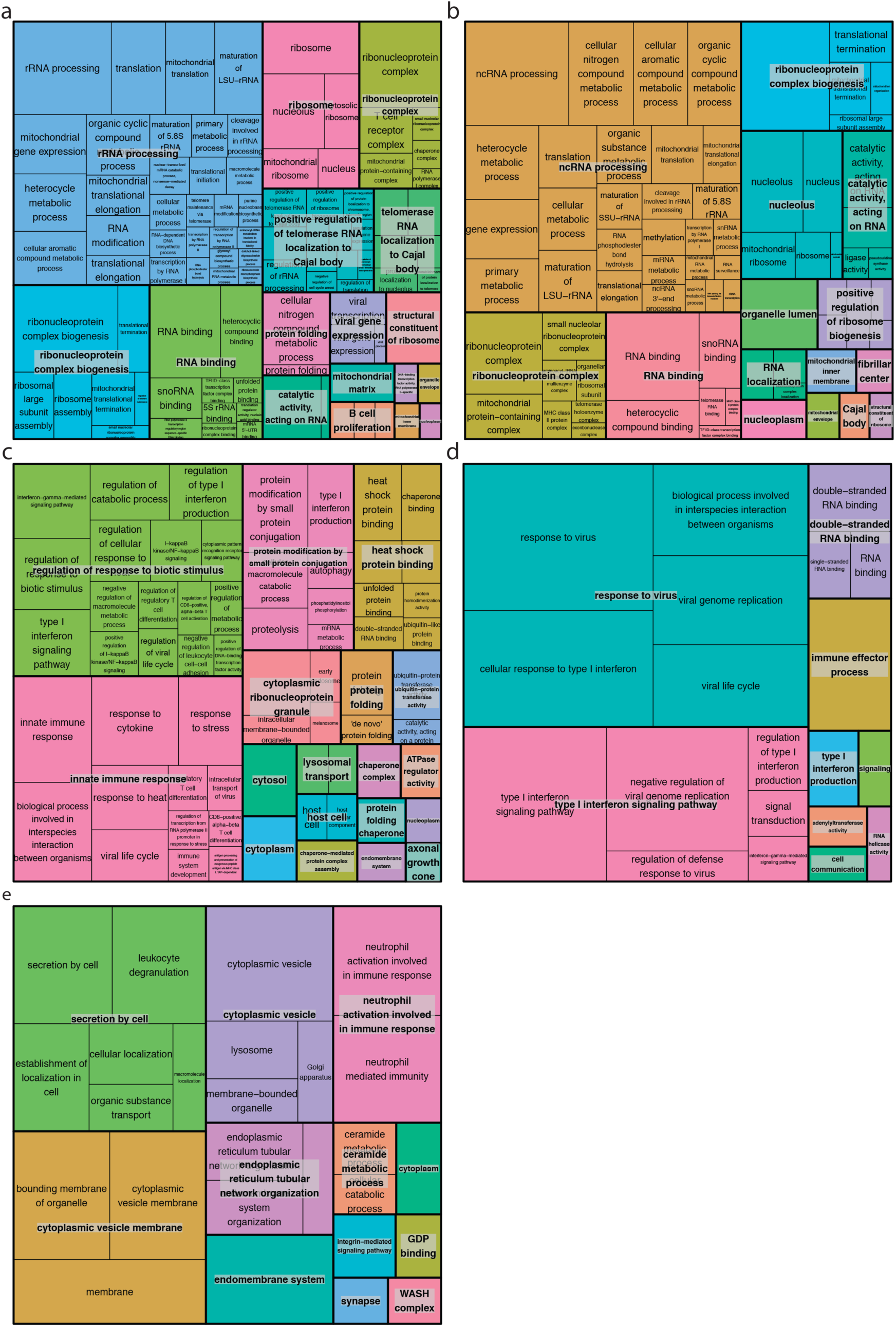
GO enrichments for DEGs for other PASC checklist items. Box sizes are relative to the -log10(adjusted p values) of the GO term enrichments for the corresponding DEGs and the term is noted in each box. Related terms are grouped by similarity and groupings are indicated by proximity and shared color. Consensus terms are indicated in bold for each group. a) Upregulated genes in memory resting CD4^+^ T cells for cavities/teeth problems. b) Downregulated genes in CD8^+^ T cells for quality of life. c) Upregulated genes in M1 macrophages for need supplemental O2. d) Upregulated genes in memory B cells for anxiety/depression. e) Upregulated genes in memory activated CD4^+^ T cells for memory/thought problems.

**Extended Data Figure 5:**
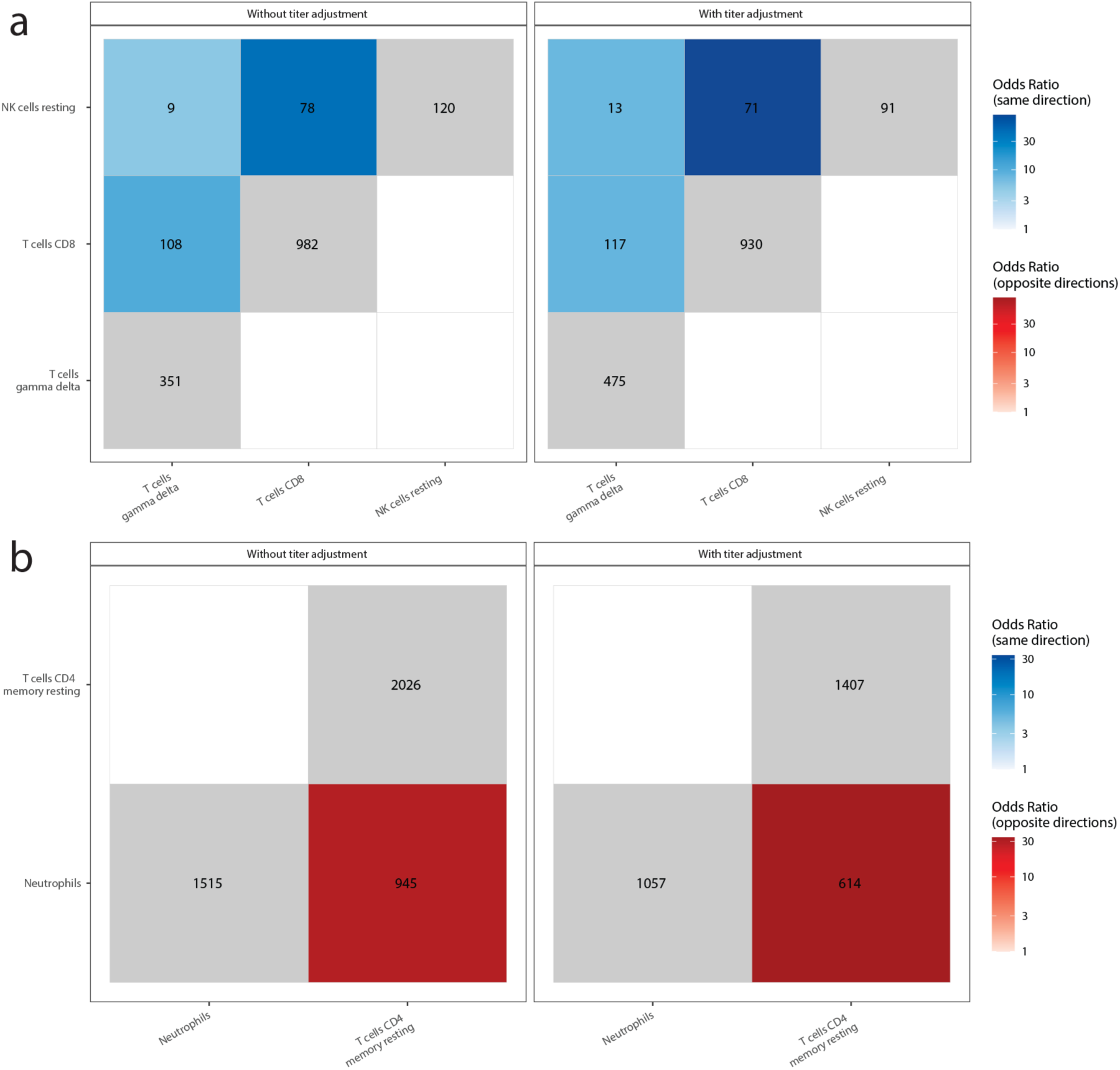
Shared PASC checklist DEGs between cell-types. The x and y axes are the cell types associated with more than 100 DEGs. The numbers in each box are the numbers of shared DEGs between the two checklist items defined in the axes, and the color represents whether they are same-direction (blue), opposite direction (red) or the total number of DEGs for that checklist item (grey). The shadings of red and blue are the ORs of the Fisher’s exact tests for the enrichment of shared DEGs in that box, and are shown only if the associated enrichment adjusted p-value < 0.05 (FWER, Holm 1979). The left and right facets represent the shared DEGs before and after adjustment for anti-spike antibody titers respectively. Symptoms in rows and columns are ordered by hierarchical clustering and optimal leaf ordering based on the shared same-direction DEGs. a) Quality of life shared DEGs. b) Cavities and teeth problems shared DEGs.

**Extended Data Figure 6:**
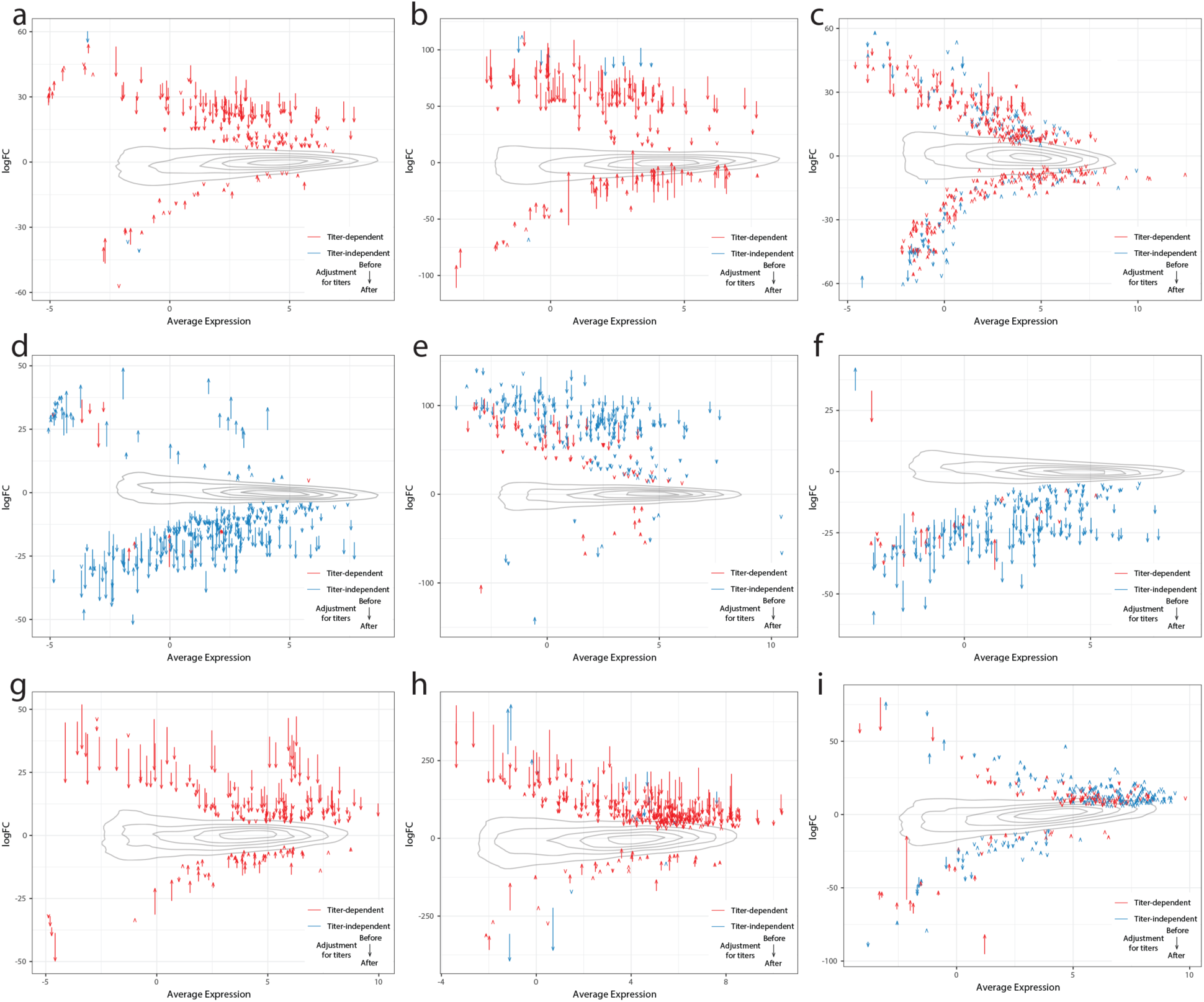
Delta-MA plot of anti-spike antibody titer effect on differential expression log2(fold change) The x and y axes represent the average normalized gene expression and the differential expression log2(fold change) respectively. Each arrow shows a single DEG. The arrow colors indicate the anti-spike antibody titer dependent (red) and independent (blue) DEGs. The contours show the distribution of all log2(fold changes) before controlling for antibody titers. The effect of controlling for antibody titers on DEGs is shown by the arrows, with the arrow tail being the log2(fold change) before adjustment and the arrow head the log2(fold change) after adjustment. a) Sleep problems in plasma cells. b) Nausea/diarrhea/vomiting in plasma cells. c) Smell/taste problems in plasma cells. d) Lung problems in plasma cells. e) Skin rash in plasma cells. f) Pneumonia in plasma cells. g) Anxiety/depression in memory B cells. h) Need Supplemental O2 in M1 macrophages. i) Memory/Thought problems in memory activated CD4^+^ T cells.

## Supplementary Data Description

### Supplementary Data 1: Cohort description and clinical associations to PASC checklist items

Description: Description of sheets and columns

Cohort Description: Population description for core cohort of 182 subjects analyzed

Symptom Prevalence: Table of prevalence of PASC symptoms in COVID-19 cases and controls Symptoms vs. Clinical Data: Table of tests for dependence of PASC symptoms on medications, labs, and comorbidities

Symptoms vs. Anti-Spike Ab: Table of tests for dependence of PASC symptoms on anti-Spike antibody titers

Symptoms vs. Cell Type Fraction: Table of tests for dependence of PASC symptoms on estimated cell type fractions

### Supplementary Data 2: Marker gene enrichments in cell-type-specific DEGs

Description: Description of sheets and columns

No Serology Adjustment: Cell type marker enrichment results for cell-type-specific DEGs with no serology adjustment

Serology Adjustment: Cell type marker enrichment results for cell-type-specific DEGs with serology adjustment

### Supplementary Data 3: PASC checklist items DE signatures before controlling for anti-spike antibody titers

Description: Description of sheets and columns

B cells memory: Table of significant DEGs (adj.P.Val <= 0.05) for B cells memory for all PASC symptoms without serology adjustment.

Macrophages M0: Table of significant DEGs (adj.P.Val <= 0.05) for Macrophages M0 for all PASC symptoms without serology adjustment.

Macrophages M1: Table of significant DEGs (adj.P.Val <= 0.05) for Macrophages M1 for all PASC symptoms without serology adjustment.

Mast cells resting: Table of significant DEGs (adj.P.Val <= 0.05) for Mast cells resting for all PASC symptoms without serology adjustment.

Monocytes: Table of significant DEGs (adj.P.Val <= 0.05) for Monocytes for all PASC symptoms without serology adjustment.

Neutrophils: Table of significant DEGs (adj.P.Val <= 0.05) for Neutrophils for all PASC symptoms without serology adjustment.

NK cells resting: Table of significant DEGs (adj.P.Val <= 0.05) for NK cells resting for all PASC symptoms without serology adjustment.

Plasma cells: Table of significant DEGs (adj.P.Val <= 0.05) for Plasma cells for all PASC symptoms without serology adjustment.

T cells CD4 memory activated: Table of significant DEGs (adj.P.Val <= 0.05) for T cells CD4 memory activated for all PASC symptoms without serology adjustment.

T cells CD4 memory resting: Table of significant DEGs (adj.P.Val <= 0.05) for T cells CD4 memory resting for all PASC symptoms without serology adjustment.

T cells CD8: Table of significant DEGs (adj.P.Val <= 0.05) for T cells CD8 for all PASC symptoms without serology adjustment.

T cells follicular helper: Table of significant DEGs (adj.P.Val <= 0.05) for T cells follicular helper for all PASC symptoms without serology adjustment.

T cells gamma delta: Table of significant DEGs (adj.P.Val <= 0.05) for T cells gamma delta for all PASC symptoms without serology adjustment.

Whole Blood: Table of significant DEGs (adj.P.Val <= 0.05) for Whole Blood for all PASC symptoms without serology adjustment.

### Supplementary Data 4: GO term enrichments for PASC checklist items DE signatures

Description: Description of sheets and columns

B cells mem. NoTA: Table of significantly enriched (adj.P.Val <= 0.05) Gene Ontology terms for B cells memory for all PASC symptoms without serology adjustment.

Macrophages M0 NoTA: Table of significantly enriched (adj.P.Val <= 0.05) Gene Ontology terms for Macrophages M0 for all PASC symptoms without serology adjustment.

Macrophages M1 NoTA: Table of significantly enriched (adj.P.Val <= 0.05) Gene Ontology terms for Macrophages M1 for all PASC symptoms without serology adjustment.

Mast cells rest NoTA: Table of significantly enriched (adj.P.Val <= 0.05) Gene Ontology terms for Mast cells resting for all PASC symptoms without serology adjustment.

Monocytes NoTA: Table of significantly enriched (adj.P.Val <= 0.05) Gene Ontology terms for Monocytes for all PASC symptoms without serology adjustment.

Neutrophils NoTA: Table of significantly enriched (adj.P.Val <= 0.05) Gene Ontology terms for Neutrophils for all PASC symptoms without serology adjustment.

NK cells rest NoTA: Table of significantly enriched (adj.P.Val <= 0.05) Gene Ontology terms for NK cells resting for all PASC symptoms without serology adjustment.

Plasma cells NoTA: Table of significantly enriched (adj.P.Val <= 0.05) Gene Ontology terms for Plasma cells for all PASC symptoms without serology adjustment.

T cells CD4 mem. act. NoTA: Table of significantly enriched (adj.P.Val <= 0.05) Gene Ontology terms for T cells CD4 memory activated for all PASC symptoms without serology adjustment.

T cells CD4 mem. rest NoTA: Table of significantly enriched (adj.P.Val <= 0.05) Gene Ontology terms for T cells CD4 memory resting for all PASC symptoms without serology adjustment.

T cells CD8 NoTA: Table of significantly enriched (adj.P.Val <= 0.05) Gene Ontology terms for T cells CD8 for all PASC symptoms without serology adjustment.

T cells gamma delta NoTA: Table of significantly enriched (adj.P.Val <= 0.05) Gene Ontology terms for T cells gamma delta for all PASC symptoms without serology adjustment.

B cells mem. TA: Table of significantly enriched (adj.P.Val <= 0.05) Gene Ontology terms for B cells memory for all PASC symptoms with serology adjustment.

Macrophages M1 TA: Table of significantly enriched (adj.P.Val <= 0.05) Gene Ontology terms for Macrophages M1 for all PASC symptoms with serology adjustment.

Monocytes TA: Table of significantly enriched (adj.P.Val <= 0.05) Gene Ontology terms for Monocytes for all PASC symptoms with serology adjustment.

Neutrophils TA: Table of significantly enriched (adj.P.Val <= 0.05) Gene Ontology terms for Neutrophils for all PASC symptoms with serology adjustment.

NK cells rest TA: Table of significantly enriched (adj.P.Val <= 0.05) Gene Ontology terms for NK cells resting for all PASC symptoms with serology adjustment.

Plasma cells TA: Table of significantly enriched (adj.P.Val <= 0.05) Gene Ontology terms for Plasma cells for all PASC symptoms with serology adjustment.

T cells CD4 mem. act. TA: Table of significantly enriched (adj.P.Val <= 0.05) Gene Ontology terms for T cells CD4 memory activated for all PASC symptoms with serology adjustment.

T cells CD4 mem. rest TA: Table of significantly enriched (adj.P.Val <= 0.05) Gene Ontology terms for T cells CD4 memory resting for all PASC symptoms with serology adjustment.

T cells CD8 TA: Table of significantly enriched (adj.P.Val <= 0.05) Gene Ontology terms for T cells CD8 for all PASC symptoms with serology adjustment.

T cells gamma delta TA: Table of significantly enriched (adj.P.Val <= 0.05) Gene Ontology terms for T cells gamma delta for all PASC symptoms with serology adjustment.

### Supplementary Data 5: PASC checklist items DE signatures while controlling for anti-spike antibody titers

Description: Description of sheets and columns

B cells memory: Table of significant DEGs (adj.P.Val <= 0.05) for B cells memory for all PASC symptoms with serology adjustment

Macrophages M0: Table of significant DEGs (adj.P.Val <= 0.05) for Macrophages M0 for all PASC symptoms with serology adjustment

Macrophages M1: Table of significant DEGs (adj.P.Val <= 0.05) for Macrophages M1 for all PASC symptoms with serology adjustment

Mast cells resting: Table of significant DEGs (adj.P.Val <= 0.05) for Mast cells resting for all PASC symptoms with serology adjustment

Monocytes: Table of significant DEGs (adj.P.Val <= 0.05) for Monocytes for all PASC symptoms with serology adjustment

Neutrophils: Table of significant DEGs (adj.P.Val <= 0.05) for Neutrophils for all PASC symptoms with serology adjustment

NK cells resting: Table of significant DEGs (adj.P.Val <= 0.05) for NK cells resting for all PASC symptoms with serology adjustment

Plasma cells: Table of significant DEGs (adj.P.Val <= 0.05) for Plasma cells for all PASC symptoms with serology adjustment

T cells CD4 memory activated: Table of significant DEGs (adj.P.Val <= 0.05) for T cells CD4 memory activated for all PASC symptoms with serology adjustment

T cells CD4 memory resting: Table of significant DEGs (adj.P.Val <= 0.05) for T cells CD4 memory resting for all PASC symptoms with serology adjustment

T cells CD8: Table of significant DEGs (adj.P.Val <= 0.05) for T cells CD8 for all PASC symptoms with serology adjustment

T cells follicular helper: Table of significant DEGs (adj.P.Val <= 0.05) for T cells follicular helper for all PASC symptoms with serology adjustment

T cells gamma delta: Table of significant DEGs (adj.P.Val <= 0.05) for T cells gamma delta for all PASC symptoms with serology adjustment

Whole Blood: Table of significant DEGs (adj.P.Val <= 0.05) for Whole Blood for all PASC symptoms with serology adjustment

### Supplementary Data 6: Cell-type mapping for marker gene enrichments

Description: Description of sheets and columns

DEG Cell Type mappings: Table of cell type mappings from LM22 reference to broad categories used to assemble lists of cell-type specific DEGs

Marker Gene Cell Type Mappings: Table of cell type mappings used to annotate cell type marker genes to broad categories from the scientific literature

